# Payments to healthcare organisations reported by the medical device industry in Europe from 2017 to 2019: an observational study

**DOI:** 10.1101/2023.04.26.23289083

**Authors:** James Larkin, Shai Mulinari, Piotr Ozieranski, Kevin Lynch, Tom Fahey, Akihiko Ozaki, Frank Moriarty

**Affiliations:** Department of General Practice, RCSI University of Medicine and Health Sciences, Dublin, Ireland; Department of Sociology, Lunds Universitet, Lund, Sweden; Department of Social and Policy Sciences, Centre for the Analysis of Social Policy, University of Bath, Bath, UK; Unaffiliated; Department of Breast and Thyroid Surgery, Jyoban Hospital of Tokiwa Foundation, Iwaki, Fukushima, Japan; Medical Governance Research Institute, Tokyo, Japan; School of Pharmacy and Biomolecular Sciences, RCSI University of Medicine and Health Sciences, Dublin, Ireland

**Keywords:** Conflicts of interest, Education, Medical device companies, Medical devices, Medical education, Payment, Pharmaceutical companies, Physician Payments Sunshine Act, Physician payment, Physician reporting, health policy, Disclosures, Transparency, Self-regulation

## Abstract

**Background:** Medical device industry payments to healthcare organisations (HCOs) can create conflicts of interest which can undermine patient care. One way of addressing this concern is by enhancing transparency of industry financial support to HCOs. MedTech Europe, a medical device trade body, operate a system of disclosure of education payments to European HCOs. This study aimed to characterise payments reported in this database and to evaluate the disclosure system.

**Methods:** An observational study of education-related payments to HCOs reported by the medical device industry in Europe was conducted. Data was manually extracted from transparentmedtech.eu. The primary outcome variable is the value of the payments, overall, and for each year, payment type, and country. The accessibility, availability and quality of the database was also analysed, using a proforma with 15 measures.

**Findings:** Overall, 116 medical device companies reported education-related payments in 53 countries, valuing over €420 million between 2017-2019, increasing in value between 2017-2019, from €91,289,672 to €175,414,302. Ten countries accounted for 94% of all payments and ten companies accounted for 80% of all payments. The accessibility, availability and quality of the database, rated low for six measures, medium for six measures and high for three measures.

**Interpretation:** There is a large amount of education-related payments from medical device companies to European HCOs, creating substantial potential for conflicts of interest. MedTech Europe’s disclosure system has many shortcomings. A European-wide publicly mandated disclosure system for both the medical device and pharmaceutical industries should be introduced.

**Funding:** Swedish Research Council (SM, PO)

## Introduction

Each year, billions of euro are paid by medical device and pharmaceutical companies to healthcare professionals (HCPs) and healthcare organisations (HCOs), ostensibly for research, consultancy and for HCPs’ education, among other areas.^1-5^ The medical device industry accounts for a large proportion of this, with some estimating that they make up the majority of industry payments.^1^ Medical devices are a vital component for health systems, used for diagnosis, treatment, and as aids to everyday activities.^6^ Despite this, the vast majority of research has been devoted to examining the patterns, nature and effects of pharmaceutical industry payments to HCOs and HCPs.^2-5^ Payments from industry to HCPs and HCOs can create conflicts of interest.^7^ Extensive evidence shows that receipt of payments from the pharmaceutical industry is associated with higher prescribing rates, higher prescribing costs, and lower prescribing quality.^8-10^ Recent research suggests that similar issues may exist for payments associated with medical devices.^11,12^ For example, a 2020 study showed that US physicians’ choice of medical device is associated with medical device industry payments.^11^ Some medical device industry payments have also been associated with legal breaches, for example of the US Anti-Kickback Statute which prohibits directly or indirectly offering or paying anything of value to induce physicians to procure certain medical products, such as devices, under state programs.^13^

To start addressing these issues, some countries, such as the United States and France, have introduced a legal requirement for disclosure of payments by industry to HCPs and HCOs.^14^ However, the preferred approach among European countries is industry self-regulation, which is based on codes of practice developed and implemented by industry trade associations, rather than legal requirements.^14^ As part of this, industry bodies have implemented self-governed systems of disclosure for payments to HCPs and HCOs; such systems are associated with a range of limitations, such as poor accessibility and limited levels of disclosure.^14^ In Europe, there is a growing amount of research on industry payments to HCPs and HCOs, but it almost exclusively focusses on the pharmaceutical industry.^4,5,8,14-17^ This study is, to our knowledge, the first European and multi-country study of medical device industry payments to healthcare organisations.

The importance of this research is highlighted by the recent implementation of the European Medical Devices Regulation, which aims to promote transparency in the medical device industry through the establishment of a comprehensive EU database on medical devices.^18^ However, when it comes to the disclosure of payments made by the medical device industry, it is MedTech Europe, the primary trade association for the European medical device industry, that oversees and regulates the European transparency.

### Policy Background

In January 2023, 137 companies were members of MedTech Europe.^19^ However, there are many medical device companies who are not members of MedTech Europe. For example, a 2015 article estimated that less than 10% of medical device companies operating in the UK were members of the Association of British Healthcare Industries, which is a member of MedTech Europe. Examples of MedTech Europe member companies include Roche, Medtronic and Abbott.^19^ Notably, several member companies have large pharmaceutical portfolios, such as Johnson and Johnson^20^ and Baxter.^21^

MedTech Europe has implemented a code of practice for their member companies to disclose details of ‘educational grants’ on a website (www.transparentmedtech.eu).^22,23^ These educational grants are described by MedTech Europe as supporting “Healthcare Professionals’ independent Medical Education”, though they also include grants for ‘public awareness campaigns’ as well as scholarships and fellowships.^22^ The MedTech Europe disclosure website, www.transparentmedtech.eu, requires member companies to report education related payments to HCOs registered in member countries of the European Free Trade Area (Switzerland, Norway, Liechtenstein and Iceland), the United Kingdom Russia, Turkey and the European Union.^23^ A Europe-wide database is in contrast to the approach of the European Federation of Pharmaceutical Industries and Associations (EFPIA) which allows each European country to set up its own disclosure system.^14^ However, it should be noted that there are exceptions for four EU countries (Belgium, France, the Netherlands and Portugal), because, in those countries, the MedTech Europe disclosure website is superseded by local laws.^23^

Publications on the transparentmedtech.eu website began in 2018, for payments made in 2017.^24^ Only payments to HCOs are reported. According to the MedTech Europe definition, HCOs include hospitals, clinics, laboratories, pharmacies, research institutions, foundations, universities and professional societies (full definition in Appendix Box 1). Notably, patient organisations are excluded from the MedTech Europe definition of HCOs. For the definition of medical devices used by MedTech Europe see Appendix Box 2. In 2018 MedTech Europe introduced a rule that member companies could no longer make payments directly to healthcare professionals for third party organised events; instead payments would be made to HCOs such as hospitals or professional societies, acting as intermediaries, who would then disburse the payments to HCPs.^22^

### Aim and objectives

Overall, a Europe-wide database offers a unique opportunity to understand the scale of payments made by the medical device industry to healthcare organisations in Europe, and to inform the development of regulation in this area. To achieve this, the aim of the study is to characterise payments reported in the *MedTech Europe* database from medical device companies to HCOs from 2017 to 2019 and to evaluate the system of disclosure itself.

## Methods

This is an observational study of payments to HCOs reported by the medical device industry in Europe. It includes annual data from 2017 to 2019 provided by medical device companies who are members of MedTech Europe. A protocol was registered for this study in February 2022.^25^ Details of deviations from the protocol are in Appendix Box 3.

### Database overview

The MedTech Europe disclosure website contains data on two categories of payments: 1) *support to educational events* and 2) *other educational grants*. Definitions are not provided for these categories but examples are. Educational events include ‘Support for HCP Participation at Third Party Organised Educational Events’ and other educational grants include ‘Scholarships, Fellowships and/or Grants for Public Awareness Campaigns’ as well as educational grants to support general medical education topics. Payments to HCOs are supposed to be aggregated on a year/donation-type basis – for example all payments under ‘other educational grants’ in 2017 by one medical device company to one HCO should be included in a single entry, though there were multiple examples of this not being adhered to

### Data extraction and processing

A data extraction form was developed in Microsoft Excel, capturing details of medical device company, year of payment, recipient identifying details (HCO name and country of registration) and payment value. Data was manually extracted from www.transparentmedtech.eu, via an exhaustive search (details in Appendix Box 4).

In some instances, medical device company subsidiaries or affiliates reported separately from their parent company on the transparentmedtech.eu website. For analysis purposes, these companies were merged (Appendix Table 1). To determine the relationship between reporting companies the following information was used: 1) The contact email address provided by the reporting entity on the website and 2) the parent company’s website and where available the subsidiary/affiliate’s website.

**Table 1.** Payments broken down by payment type

|  | 2017 | 2018 | 2019 | Total |
| --- | --- | --- | --- | --- |
| *Overall spending (% of total)* |  |  |  |  |
| Support to Educational Events | €77,428,198 (84.8%) | €122,768,363 (79.8%) | €132,400,996 (75.5%) | €332,597,557 (79.1%) |
| Other Educational Grants | €13,861,474 (15.2%) | €31,088,294 (20.2%) | €43,013,306 (24.5%) | €87,963,074 (20.9%) |
| Total | €91,289,672 | €153,856,657 | €175,414,302 | €420,560,631 |
| *Value (€) per entry, mean (SD)* |  |  |  |  |
| Support to Educational Events | 20,116 (694,978) | 14,676 (544,128) | 15,635 (541,871) | 16,082 (574,305) |
| Other Educational Grants | 15,557 (27,334) | 23,659 (85,867) | 19,288 (148,074) | 19,834 (115,602) |
| Overall | 19,259 (626,361) | 15,896 (506,839) | 16,397 (486,809) | 16,744 (523,399) |
| *Value (€) per entry, median (IQR)* |  |  |  |  |
| Support to Educational Events | 2,349 (1,000-6140) | 2,008 (888-6,140) | 2,000 (810-5,320) | 2,013 (870-5,502) |
| Other Educational Grants | 4,900 (1,495-20,000) | 5,355 (1,656-21,784) | 2,579 (733-11,000) | 3,676 (1,038-16,000) |
| Overall | 2,500 (1,000-7,898) | 2,263 (948-6,292) | 2,039 (800-6,000) | 2,238 (900-6,500) |
| *Number of unique entries (% of total)* |  |  |  |  |
| Support to Educational Events | 3,849 (81.2%) | 8,365 (86.4%) | 8,468 (79.2%) | 20,682 (82.3%) |
| Other Educational Grants | 891 (18.8%) | 1,314 (13.6%) | 2,230 (20.8%) | 4,435 (17.7%) |
| Overall | 4,740 | 9,679 | 10,698 | 25,117 |

All payments were exchanged to euro using the average annual exchange rates for the respective year published by the European Central Bank.^26^ The euro value of all totals were rounded to the nearest euro. MedTech Europe stipulate that all payments should be exclusive of VAT.^22^ Three entries totalling €14,000 were removed from 2017 for ‘Other Educational Grants’ because the recipient was ‘TEST’.

### Outcome variables

The primary outcome variable is the value of the payments, overall, and for each year, payment type, and country. A range of secondary outcomes are also included. Number of unique entries was also analysed for each year, payment type, and country. Gini coefficients are used as a measure of concentration of payments across all countries and companies. A Gini coefficient is a measure of inequality that can range from 0-1 where 1 would represent perfect inequality (e.g. one company accounting for the full value of all payments) and 0 (all companies making the same value of payments) would represent perfect equality.

The ten countries, companies and HCOs with the highest total payment value accounted for a significant proportion of the overall total value of payments. Therefore, additional secondary outcome variables are included for them. Gini coefficients are used across countries within each of the top 10 companies and across medical device companies within each of the top 10 countries. For each of the top 10 countries, companies and HCOs, the percentage of the total value of payments accounted for by each country, company or HCO was included. For each of the top ten countries, outcome variables also include: the total value of payments per 1,000 population, and the number of medical device companies reporting payments in that country. Population figures for payments per 1,000 population and Gini coefficient calculations were gathered for 2019 only, from Eurostat^27^ and World Bank data.^28^ For Gini coefficients, population figures are needed in order to understand what would have been a proportional payment value for a country. For each of the top ten companies, outcome variables also include the number of countries where payment recipients are located. For the top ten HCOs, outcome variables also include: number of companies each HCO was in receipt of payments from, the company with the greatest value of payments to each HCO, and the country the HCO is registered in.

Background information of the 10 medical device companies who made the highest total value of payments are provided in Appendix Table 2. This includes details of the medical device areas in which each company makes the greatest amount of revenue, the overall revenue of the companies, and their research and development expenditure. Information was gathered using each company’s annual report for 2019 and the MedTech Europe website.^29^

**Table 2.** Ten countries with highest value of reported payments to healthcare organisations between 2017-2019

|  | Country | Support to Educational Events | Other Educational Grants | Total value of payments (€) | € per 1,000 population | % of the total value of payments accounted for by each country | Number of medical device companies reporting payments | Gini Coefficient for Companies within Country* |
| --- | --- | --- | --- | --- | --- | --- | --- | --- |
| 1 | Switzerland | 165,872,384 | 10,623,301 | 176,495,685 | 20,661 | 42.0 | 54 | 0.96 |
| 2 | Spain | 61,850,963 | 23,317,944 | 85,168,908 | 1,815 | 20.3 | 82 | 0.78 |
| 3 | UK | 18,695,575 | 17,860,467 | 36,556,042 | 549 | 8.7 | 67 | 0.82 |
| 4 | Italy | 24,302,166 | 6,172,585 | 30,474,751 | 505 | 7.2 | 57 | 0.74 |
| 5 | Netherlands | 7,346,538 | 14,125,358 | 21,471,896 | 1,242 | 5.1 | 38 | 0.83 |
| 6 | Germany | 16,712,991 | 4,310,364 | 21,023,355 | 253 | 5.0 | 56 | 0.79 |
| 7 | Austria | 6,104,074 | 3,044,929 | 9,149,003 | 1,033 | 2.2 | 47 | 0.80 |
| 8 | Poland | 5,007,723 | 539,482 | 5,547,206 | 146 | 1.3 | 36 | 0.75 |
| 9 | Turkey | 3,436,760 | 1,058,739 | 4,495,499 | 55 | 1.1 | 20 | 0.64 |
| 10 | Russia | 2,742,170 | 1,571,552 | 4,313,722 | 30 | 1.0 | 18 | 0.61 |

The accessibility, availability and quality of the database is also included as a secondary outcome. This was assessed using an adapted version of the proforma developed by Ozieranski and colleagues to examine the accessibility and quality of pharmaceutical industry payment data.^14^ Several changes were made to the proforma to provide a more comprehensive assessment of the disclosure system (details of the changes can be seen in Appendix Box 5). The proforma contains 15 measures, nine measures of accessibility, three measures of availability and three measures of quality. The measures can be rated at one of three levels from low to high, for seven of the measures there are only two levels, low and high. The full proforma can be seen in Appendix Table 3. Other issues with the database that were not covered in the proforma assessment were inductively assessed, with further details of analysis methods detailed below.

**Table 3.** Ten medical device companies with highest value of payments disclosed between 2017-2019

|  | Company | Support to Educational Events | Other Educational Grants | Total value of payments (€) | % of the total value of payments accounted for by each company | Number of countries where payment recipients are located | Gini Coefficient for Countries within Companies* |
| --- | --- | --- | --- | --- | --- | --- | --- |
| 1 | Johnson & Johnson Medical | 171,370,841 | 11,318,575 | 182,689,416 | 43.4 | 27<br>(26 eligible) | 0.90 |
| 2 | Abbott Laboratories | 30,348,508 | 13,293,670 | 43,642,178 | 10.4 | 31<br>(24 eligible) | 0.46 |
| 3 | Boston Scientific | 19,767,471 | 13,369,855 | 33,137,326 | 7.9 | 26<br>(20 eligible) | 0.52 |
| 4 | Medtronic International Trading Sàrl | 18,160,048 | 5,594,744 | 23,754,792 | 5.6 | 29<br>(26 eligible) | 0.58 |
| 5 | Baxter | 6,807,381 | 9,220,650 | 16,028,031 | 3.8 | 23<br>(21 eligible) | 0.82 |
| 6 | Carl Zeiss Meditec Iberia | 7,998,571 | 0 | 7,998,571 | 1.9 | 1<br>(1 eligible) | N.A. |
| 7 | Smith & Nephew Orthopaedics AG | 3,291,518 | 4,304,874 | 7,596,393 | 1.8 | 31<br>(15 eligible) | 0.25 |
| 8 | BIOTRONIK | 4,082,339 | 3,403,986 | 7,486,324 | 1.8 | 18<br>(14 eligible) | 0.68 |
| 9 | Roche | 4,169,433 | 3,198,437 | 7,367,869 | 1.8 | 27<br>(24 eligible) | 0.20 |
| 10 | Zimmer GmbH | 1,934,888 | 5,036,747 | 6,971,635 | 1.7 | 20<br>(17 eligible) | 0.59 |

### Quantitative analysis

Payment values are summarised using totals, means and standard deviations, and medians and interquartile ranges. Number of unique entries is summarised using totals.

For the calculation of Gini coefficients for countries overall and for the countries within each of the top 10 companies, only MedTech Europe member countries who do not have national laws that supersede industry body guidance were included.

For analysis of HCOs, naming was inconsistent, so detailed analysis was not possible. Analysis was conducted for the top 10 HCOs only, based on of their names as reported. Quantitative analysis was conducted using R-4.1.1 software.

### Content Analysis

For analysis of the accessibility, availability and quality of the database, data was coded by one author (JL) and cross-checked by a second author (FM). During the process of data extraction, analysis and database assessment, a content analysis was conducted to document any other issues with the database that had not been included in the assessment of the accessibility, availability and quality of the database. This was conducted by one author (JL) and cross-checked by a second (SM or FM).

## Results

### Payment Patterns Overall

In total, 116 medical device companies reported payments valuing €420,560,631 between 2017-2019. Increasing from €91,289,672 in 2017 to €175,414,302 in 2019, a 92% increase. The number of companies reporting payments in each year also increased, from 66 in 2017 to 101 in 2018 and 94 in 2019 a 42% increase between 2017 and 2019. The dominant payment category in each year was *Support to Educational Events* (between 75.5% and 84.8% annually). However, *Other Educational Grants* showed a higher overall increase (15.2% of all payments in 2017 vs 24.5% in 2019). More details can be found in table 1.

### Countries

In total, payments were reported in 53 countries (Appendix Tables 4 & 5). Notably, payments were reported in the four countries with superseding national legislation; €21,471,896 in the Netherlands, €119,387 in Portugal, €2,176,609 in France and €1,517,567 in Belgium (Appendix Table 4). Also, €431,623 in payments was reported in 28 countries that were not members of MedTech Europe (Appendix Table 4).

**Table 4.** Ten named recipients with highest value of payments received between 2017-2019^*^

|  | Healthcare organisation | Support to Educational Events | Other Educational Grants | Total value of payments (€) | Number of companies in receipt of payments from | Company with greatest value of payments to HCO | % of the total value of payments accounted for by each company | Country |
| --- | --- | --- | --- | --- | --- | --- | --- | --- |
| 1 | AO Foundation** | 151,486,352 | 0 | 151,486,352 | 1 | Johnson & Johnson Medical | 36.0 | Switzerland |
| 2 | Sociedad Española De Enfermería Nefrológica | 43,000 | 6,651,587 | 6,694,587 | 1 | Baxter | 1.6 | Spain |
| 3 | European HCO and PCO | 2,847,655 | 0 | 2,847,655 | 1 | B. Braun | 0.7 | Germany |
| 4 | Fundacion para la Investigacion en Gastroenterologica y Hepatologia | 12,000 | 2,479,520 | 2,491,520 | 1 | Boston Scientific | 0.6 | Spain |
| 5 | Osteosynthesis and Trauma Care Foundation | 2,349,603 | 0 | 2,349,603 | 1 | Stryker | 0.6 | Switzerland |
| 6 | Nubbecas Focused Solutions For Companies | 2,087,273 | 0 | 2,087,273 | 1 | Baxter | 0.5 | Spain |
| 7 | CME4U GMBH | 1,991,401 | 30,376 | 2,021,777 | 6 | Boston Scientific | 0.5 | Germany |
| 8 | Fundacion Fidis | 1,591,598 | 6,700 | 1,598,298 | 1 | Baxter | 0.4 | Spain |
| 9 | Fundacion Senefro | 259,056 | 1,307,822 | 1,566,878 | 4 | Baxter | 0.3 | Spain |
| 10 | Avoris Retail Division S.L. | 1,407,407 | 0 | 1,407,407 | 3 | Coloplast | 0.3 | Spain |

However, 10 countries made up 93.8% of the total value of payments (Table 2). Switzerland made up 42.0% of the total value of payments, followed by Spain (20.3%). Notably, there is a very high concentration of payments in Switzerland, where across the 54 medical device companies making payments, the Gini coefficient was 0.96, compared to 0.78 for Spain. The Gini coefficient for the 28 countries without superseding national legislation was 0.72. When examining euros paid per 1,000 population, Switzerland remains by far the highest recipient (€20,661). One other notable country with very high levels of euros paid per 1,000 population is Luxembourg with €2,148 (Appendix Table 5). Croatia, Slovenia and Ireland also have high levels of euros paid per 1,000 population, all with over €400 per 1,000 population (Appendix Table 5). The three countries with the lowest levels of euros paid per 1,000 population were Cyprus, Bulgaria and Romania, all with less than €25 per 1,000 population (Appendix Table 5).

**Figure 1.**
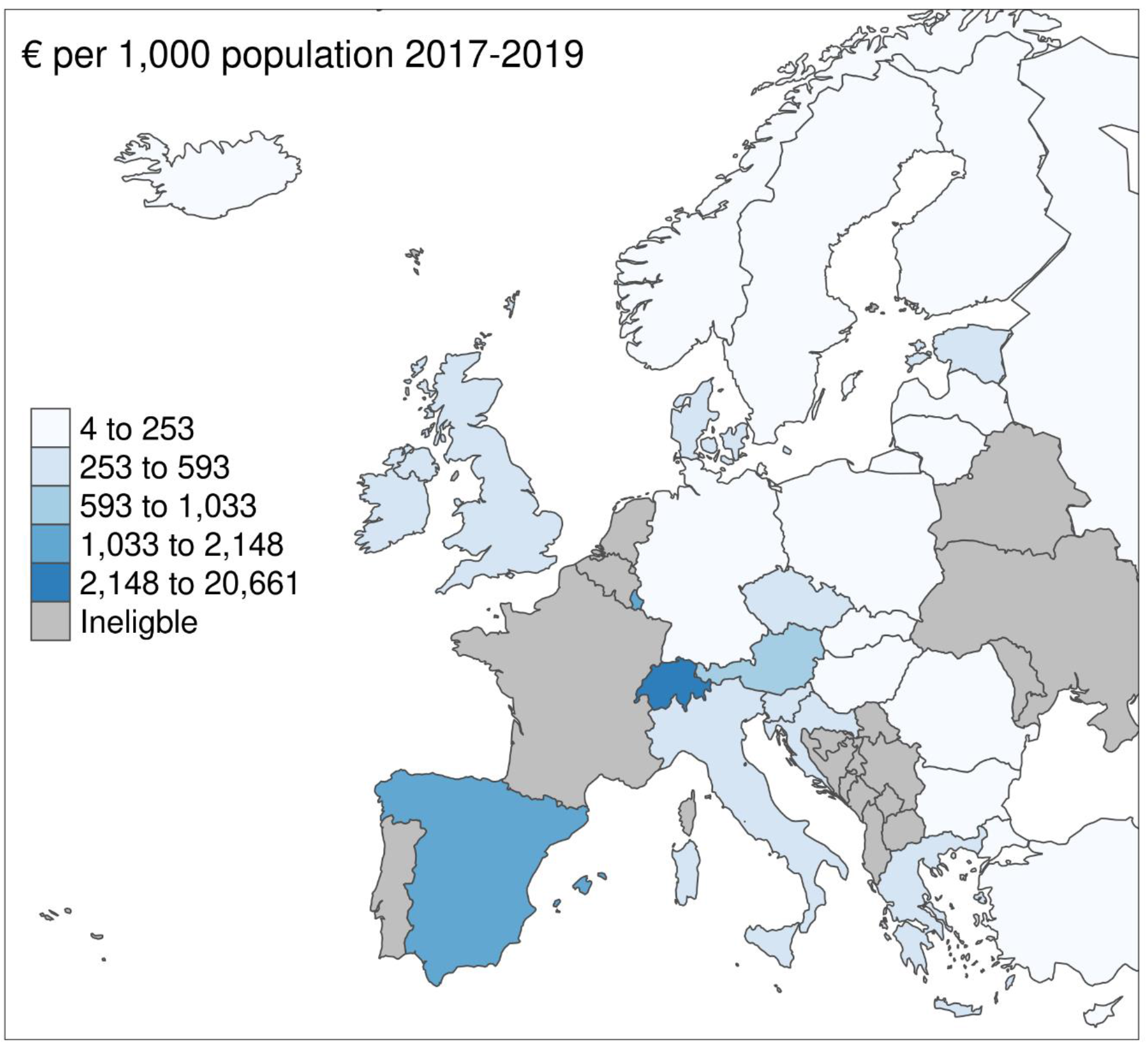
Payments (€) per 1,000 population across member countries without superseding laws from 2017 to 2019.

### Companies

Of the 116 medical device companies who reported payments between 2017-2019 (Appendix Table 1), ten companies made up 80.0% of payments (Table 3). The overall Gini coefficient for the concentration of medical device company payments to HCOs was 0.89. This high degree of concentration was largely driven by one medical device company, Johnson & Johnson Medical, who accounted for 43.4% of all payments. Johnson & Johnson Medical themselves also had a very high concentration of payments across countries; across the 26 member countries without superseding legislation, the Gini coefficient was 0.90, explained by its large investment in Switzerland, compared to 0.46 for Abbott Laboratories. Another notable element of the top 10 companies is that Medtronic International Trading Sàrl only disclosed payments in 2017 and 2018.

With regard to the type of companies in the top 10, two are involved in in-vitro diagnostic medical devices and medical devices (refer to Appendix Box 2 for distinction), and 7 are involved in medical devices only. Considering the top three product areas per company by revenue, three of top ten had cardiac rhythm management, three had orthopaedics and three had surgery. Full overviews of the top ten companies are provided in Appendix Table 2.

### Healthcare organisations

There were 13,916 uniquely named HCOs who received at least one payment between 2017 and 2019. However, this likely includes many cases of the same HCO being included with a different spelling or a different name. The top 10 HCOs are shown in Table 4. AO Technology in Switzerland was in receipt of 36% of all payments, representing approximately €151.5 million, all of which was from Johnson and Johnson Medical, for *Support to Educational Events*. Another notable entry is ‘European HCO and PCO’ which is in receipt of the third largest amount. However, this is likely to be an aggregation of several payments to different HCOs and professional conference organisers by B. Braun. Notably, seven of the top ten recipients each received all of their payments from single companies. Also, of note, six of the top ten companies are registered in Spain and the other four are registered in either Germany or Switzerland.

### Accessibility, Availability and Quality of the Database

Overall, using the 15 measures of accessibility, availability and quality, the database rated low on six measures, medium on six measures and high on three measures. The three areas where the database rated high were 1) the database format was a website as opposed to individual PDFs, 2) structure; all companies follow a single template, and 3) clear tax reporting; VAT is excluded for all entries. The six areas where the database rated low were 1) availability of customisable summary statistics, 2) downloadability, 3) removal of data after certain number of years, 4) breadth of recipients, 5) breadth of donors, and 6) payment areas did not include research, consulting, gifts, and charitable donations. For more details see Appendix Table 3. Other issues with the database that were inductively noted included 1) itemisation of payments, contrary to guidance^22^ 2) disclosure of payments to patient organisations and individual HCPs, contrary to guidance^22^ and 3) instances of payments disclosed on the transparentmedtech.eu website but not the respective mandatory disclosure system. Full details are included in Appendix Box 6.

## Discussion

### Summary

Between 2017 and 2019 medical device companies disclosed €420 million in educational grants to healthcare organisations creating a large potential for conflicts of interest. These payments were made by 116 medical device companies to HCOs registered in 53 countries. Eighty percent of payments were accounted for by the top 10 companies, and Johnson and Johnson Medical accounted for 43% of payments which were primarily to one healthcare organisation, AO Foundation in Switzerland. Seventy-nine percent of the €420 million in educational grants were for HCP participation at third party organised educational events, and the other 21% was for scholarships, fellowships, educational grants to support general medical education topics or for public awareness campaigns. The database was rated as low for six measures of accessibility, availability and quality, medium for six measures and high for three measures, indicating major shortcomings with this system of disclosure.

### Results in Context

Unlike in the legally-mandated US Open Payments database, no information is provided on the therapeutic area or device or test that each payment relates to. However, the examination of the top recipients, who account for 42% of all payments, provides an opportunity to understand what areas the payments relate to. AO foundation is the largest recipient. This HCO hosts educational events for surgeons, in the areas of ‘trauma and disorders of the musculoskeletal system.’^30^ The fifth highest recipient, Osteosynthesis and Trauma Care Foundation, is also a HCO with a focus on orthopaedics.^31^ Two of the top ten are nephrology HCOs. However, several of the top ten HCOs appear to be event organisers or travel companies (e.g. Nubbecas Focused Solutions For Companies,^32^ CME4U GMBH,^33^ Avoris Retail Division S.L.^34^) making it difficult to know what healthcare areas their payments relate to. In terms of product areas that generated the most revenue for the top ten medical device companies: cardiology, orthopaedics, and surgery appeared the most frequently. These areas are similar to four of the top five payment areas in the US for medical devices.^1^

In terms of the countries HCOs are registered in, two of the top ten are based in Switzerland; AO Foundation and Osteosynthesis and Trauma Care Foundation. However, both organisations appear to hold events in multiple countries.^30,31^ If payments registered for Switzerland are actually used in other countries, it partly undermines the validity of the database. Six of the top ten companies are registered in Spain, and for at least three of these companies,^35-37^ their activities appear to be primarily in Spain. Given this information, along with the fact that Spain is one of the highest recipients of payments, it implies that industry marketing activities might be especially prominent there. This is reinforced by a recent study of the pharmaceutical industry which found that Spain stood out in a seven-country comparison because of its higher payment amounts.^4^ There are a few countries with relatively low payments, like Romania, Cyprus and Bulgaria. The reasons for such large disparities between countries are unclear but may include, among other reasons, culture, demographics, regulations, the organisation of healthcare and national association compliance.^38^ Further research is needed to elucidate these issues.

Also of note is the large amount of payments reported in countries with separate mandatory reporting systems (Belgium, France, Netherlands and Portugal). For example, €21 million in payments were declared in the Netherlands. This creates confusion as it is unclear whether these payments were or were not also reported via the national mandatory disclosure systems. As noted in the content analysis, there are several cases of payments disclosed on the transparentmedtech.eu website but not the respective mandatory disclosure system.

In terms of the concentration of payments, there was a similar Gini coefficient for UK pharmaceutical companies; 0.85^5^ compared to 0.89 for European medical device companies, demonstrating the dominance of a few companies. Also, the large concentration of payments amongst the top ten medical device companies is similar to the top ten pharmaceutical companies making payments to HCOs in the UK. In the UK, the top ten companies made up 82% of the total value of payments,^5^ compared to 80% for medical device companies in Europe.

Overall, the figure of €420 million likely underestimates the true extent of medical device industry payments. Firstly, the large increase in payments between 2017 and 2018 is very likely a function of the rule introduced in 2018 that member companies could no longer make payments directly to healthcare professionals for attendance at third party organised educational events. Instead payments would be made to HCOs such as hospitals or professional societies, acting as intermediaries, who would then make the payments to healthcare professionals.^22^ Therefore, the 2017 figure is likely a large underestimate of the value of educational grants provided. Secondly, it should be noted that according to MedTech Europe, in 2019, 5% of MedTech Europe national associations had not banned their member companies from providing direct sponsorship of healthcare professionals.^38^ So the figures for 2018 and 2019 are also likely to be underestimates.

Thirdly, it is not clear whether all medical device companies are reporting all relevant payments. For example, Medtronic reported no payments in 2019 despite disclosing payments of almost €24 million across 2017 and 2018. This may be as a result of no payments being made or alternatively no disclosure being submitted to transparentmedtech.eu. Like self-regulatory codes for the pharmaceutical industry,^17^ MedTech Europe does not appear to require member companies to make a report if they have not made payment in a given year. Under-reporting has been documented in other self-regulatory systems.^15,16^ Finally, many companies are not members of MedTech Europe or the national associations within MedTech Europe,^39^ which is likely to also lead to large underestimates of payments. This is a consistent problem with systems that employ self-regulation.^17^

In terms of accessibility and quality of the database, its general low rating was similar to the rating of the few existing pharmaceutical industry disclosure databases in European countries, most of which are also not downloadable, do not use unique identifiers consistently, and do not make customisable summary statistics available.^14^ The two areas that transparentmedtech.eu scored medium or high, where most pharmaceutical industry disclosure systems scored low,^14^ were availability of a limited search function and clarity of tax inclusion, respectively.

A major limitation is the narrow breadth of payments covered by the database, far more limited than most pharmaceutical industry databases, be they publicly mandated or self-regulatory.^14^ It should be noted that Eucomed, a representative body for European medical device companies which is now part of MedTech Europe,^40^ previously considered disclosing payments that covered areas other than education, these included 1) Consultancy fees and expenses related to consultancy, 2) Charitable donations, 3) Research grants, and 4) Gifts and give-aways.^41^ Also, many pharmaceutical industry disclosure websites disclose research and development payments to healthcare professionals and healthcare organisations, and these make up a large proportion of payments (though in Europe this is usually presented as an aggregated value for each company).^42^ Nonetheless, these areas are not covered by the transparentmedtech.eu database.^22^ Another major area that is not covered by transparentmedtech.eu, but is covered by US Open Payments, is ownership or shares in medical device companies.^22,43^ Wider coverage of payments is very important, as several of these non-covered areas (e.g. consulting payments) have been found, on occasion, to be used for improper payments to physicians.^44^ The limited breadth of payment areas along with the other shortcomings discussed above may be the primary reason for the large differences between the payment levels found in this research, compared to those found in studies of the UK pharmaceutical industry and the US medical device industry. Pharmaceutical companies reporting payments on the self-regulatory disclosure website for the British pharmaceutical industry^45^ made payments to HCOs and HCPs registered in the UK valuing €2.96 billion between 2017 and 2019,^46^ compared to the €36.6 *million* reported by medical device companies to UK HCOs on transparentmedtech.eu. In the US, medical device companies pay about $904 million annually, just to physicians.^1^

Overall, the usefulness of the database is severely limited, and it does not provide transparency on the true scale of payments. Given the deficiencies, there does not appear to be a clear intended user or audience for the database. Like other self-regulated disclosure systems, it may instead be a means of creating an appearance of credibility and compliance in order to avoid regulation of the industry’s marketing activities.^47^

### Strengths and limitations

A major strength of this study is the large number of countries, companies covered, 53 and 116 respectively. Also, this is the first Europe-wide analysis of medical device industry payments to HCOs. Another strength of this study was the analysis of data over a three-year period. A further strength of this study is the advancement of the database assessment tool developed by Ozieranski and colleagues.^14^

A limitation is the lack of analysis of HCOs. This was due to the inconsistent use of unique identifiers by medical device companies when reporting payments. Also, the inability to download the database meant that manual data extraction was conducted which may have led to errors. Many of the other limitations with the study are associated with the accessibility, quality and availability of the data itself, discussed above.

### Implications

This research adds to the extensive literature documenting the shortcomings of self-regulated disclosure systems for industry payments to HCPs and HCOs.^2,3,5,14,48^ Overall the shortcomings of this database are reflective of general issues seen with self-regulation across several industries, such as pharmaceutical,^49,50^ nutrition^51^ and alcohol.^52^ This highlights the need for a publicly mandated database. This could be EU wide and cover both the medical device and pharmaceutical industry.

Several groups have suggested harmonisation of minimum standards for transparency across Europe, which would include a high-quality pan-European database.^14,17^ The database could be downloadable, cover a broad range of payments to healthcare professionals, HCOs and patient organisations, and provide information on the therapeutic area and device or test that each payment relates to in ways that allow integration with the planned EU database on medical devices.^18^

The main implication of this study is the large potential for conflicts of interest in European healthcare arising from the large value of the payments (at least €420 million) being made by the medical device industry to HCOs related to education. These payments provide medical device companies with an opportunity to influence a range of HCOs such as hospitals, universities, clinical societies and professional training bodies, all of which have a significant influence on healthcare practice. There is extensive evidence suggesting that payments of this nature have a negative influence on pharmaceutical prescribing.^8-10^ There is some evidence that these payments are associated with sub-optimal medical device procurement in the USA,^11,12^ though more research is needed in this area to understand the relationship between medical device industry interaction and clinical practice across countries.

As pointed out in a seminal US Institute of Medicine report: ‘The disclosure of individual and institutional financial relationships is a critical but limited first step in the process of identifying and responding to conflicts of interest.’^7^ Many have called for greater restrictions in the relationship between industry and physicians/HCOs.^7,53,54^ However, restrictions around education may create a funding gap. Alternative funding sources such as state funding, a hypothecated medica device industry tax,^49^ or HCPs paying for education using their own personal income, could be considered.

## Conclusion

This study shows the large amount of payments made by the medical device industry to healthcare organisations in 53 countries, primarily in Europe. While this provides a first estimate of the scale of payments, the major shortcomings with the database and the reporting requirements make it likely that the total value of payments is significantly larger. An EU mandated system of disclosure for the medical device and pharmaceutical industries could address these shortcomings, and enhance transparency in the healthcare sector’s interactions with industry. However, greater transparency is just one step in addressing the potential negative effects that industry payments to healthcare organisations can have on healthcare.

## Data Availability

Aggregated data are available online at:
Larkin, James, Lynch, Kevin, & Moriarty, Frank. (2023). Medical Device Industry Payments Europe 2017-2019 [Data set]. Zenodo. https://doi.org/10.5281/zenodo.7866632

## CREDIT Statement

James Larkin: conceptualisation, data curation, formal analysis, methodology, writing – original draft

Shai Mulinari: formal analysis, methodology, writing – review & editing

Piotr Ozieranski: methodology, writing – review & editing.

Kevin Lynch: investigation, writing – review & editing.

Tom Fahey: conceptualisation, writing – review & editing.

Akihiko Ozaki: writing – review & editing.

Frank Moriarty: conceptualisation, formal analysis, visualisation, writing – review & editing.

