## Appendix A - Supplementary Data for "Payments to healthcare organisations reported by the medical device industry in Europe from 2017 to 2019: an observational study"

### Box 1. MedTech Europe definition of healthcare organisation

Defined by MedTech Europe as any legal entity or body (irrespective of its legal or organisational form) that is a healthcare, medical or scientific association or organisation which may have a direct or indirect influence on the prescription, recommendation, purchase, order, supply, utilisation, sale or lease of medical technologies or related services such as a hospital or group purchasing organisation, clinic, laboratory, pharmacy, research institution, foundation, university or other teaching institution or learned or professional society (except for patient organisations); or through which one or more Healthcare Professionals provide services.

### Box 2. Medical device and in vitro diagnostic medical device definitions according to EU regulations

**Medical device definition^1^**

medical device’ means any instrument, apparatus, appliance, software, implant, reagent, material or other article intended by the manufacturer to be used, alone or in combination, for human beings for one or more of the following specific medical purposes:

— diagnosis, prevention, monitoring, prediction, prognosis, treatment or alleviation of disease,

— diagnosis, monitoring, treatment, alleviation of, or compensation for, an injury or disability, — investigation, replacement or modification of the anatomy or of a physiological or pathological process or state,

— providing information by means of in vitro examination of specimens derived from the human body, including organ, blood and tissue donations,

and which does not achieve its principal intended action by pharmacological, immunological or metabolic means, in or on the human body, but which may be assisted in its function by such means. The following products shall also be deemed to be medical devices:

— devices for the control or support of conception;

— products specifically intended for the cleaning, disinfection or sterilisation of devices as referred to in Article 1(4) and of those referred to in the first paragraph of this point.

**In vitro medical device^2^**

‘in vitro diagnostic medical device’ means any medical device which is a reagent, reagent product, calibrator, control material, kit, instrument, apparatus, piece of equipment, software or system, whether used alone or in combination, intended by the manufacturer to be used in vitro for the examination of specimens, including blood and tissue donations, derived from the human body, solely or principally for the purpose of providing information

on one or more of the following:

(a) concerning a physiological or pathological process or state;

(b) concerning congenital physical or mental impairments;

(c) concerning the predisposition to a medical condition or a disease;

(d) to determine the safety and compatibility with potential recipients;

(e) to predict treatment response or reactions;

(f) to define or monitoring therapeutic measures.

Specimen receptacles shall also be deemed to be in vitro diagnostic medical devices;

### Box 3. Deviations from the protocol

The following was planned in the protocol but not completed in the study.

- We will also detail the proportional value of the top 10% and bottom 75% of payments at the donor and payment level.
- The Gini coefficients illustrated using a Lorenz curve.
- Emailing each company requesting their respective methodology note for 2019
- Email MedTech Europe a modified version of the questionnaire for industry trade groups developed by Ozieranski and colleagues

The following was not outlined in the protocol but conducted in the study

- Use of R-4.1.1 software
- Addition of areas to accessibility, availability and quality proforma outlined in Box 4 other than methodology note measure which was outlined in the protocol
- Details of manual extraction process
- Detailing the 10 medical device companies who made the highest total value of payments.
- Detailing the 10 countries with highest values of payments received by HCOs
- Detailing the 10 HCOs, in receipt of the highest values of payments
- The inductive content analysis examining other issues in terms of accessibility availability and quality of the database.

### Box 4. Full description of the manual data extraction process

Manual data extraction involved entering one letter of the alphabet (e.g. b) into the search bar. Then data was copied and pasted into the data extraction sheet webpage by webpage – each page had 10 results only. For each search, data extraction was only conducted for HCOs beginning with the letter searched, to avoid duplication. A maximum of 100 pages displayed for a search. In cases where all HCOs beginning with the searched letter (e.g. a) were not included in the 100 pages, an extended search was conducted for that letter (e.g. aa, ab, ac etc.). To gather data on the country the HCO was registered in, it was necessary to click into an entry related to each HCO and then enter the data into the excel file.

### Table 1 Payments broken down by medical device company

| **Companies** | **Total value of payments (€)** | **Companies merged** | **Total value of payments (€)** |
| --- | --- | --- | --- |
| 3M | 258,343 | 3M | 507,819 |
| 3M ESPAÑA SL | 249,477 |  |  |
| Abbott Laboratories | 43,642,178 | Abbott Laboratories | 43,642,178 |
| Acelity | 24,568 | Acelity | 24,568 |
| Acumed Iberica | 123,485 | Acumed Iberica | 123,485 |
| Aesculap Chifa Sp. z o.o. | 1,974 | Aesculap Chifa Sp. z o.o. | 1,974 |
| Air Liquide Healthcare Ireland | 2,100 | Air Liquide Healthcare Ireland | 2,100 |
| Alcon | 2,356,348 | ALCON | 3,799,029 |
| ALCON BULGARIA EOOD | 6,136 |  |  |
| ALCON CUSÍ, S.A | 439,266 |  |  |
| ALCON EYE CARE UK LTD | 234,444 |  |  |
| Alcon Hungária Kft. | 8,546 |  |  |
| ALCON ITALIA S.P.A. | 202,571 |  |  |
| ALCON LABORATORIES HELLAS COMMERCIAL & INDUSTRIAL S.A. | 20,000 |  |  |
| ALCON LABORATUVARLARI TICARET A.S | 2,128 |  |  |
| ALCON NEDERLAND b.v. | 14,821 |  |  |
| ALCON NORDICS A/S - Denmark | 13,417 |  |  |
| ALCON OPHTHALMIKA GmbH | 2663 |  |  |
| ALCON PHARMACEUTICALS (Czech Republic) s.r.o | 70251 |  |  |
| ALCON PHARMACEUTICALS LTD. | 17412 |  |  |
| ALCON PHARMACEUTICALS Ltd. BRANCH OFFICE | 35600 |  |  |
| ALCON POLSKA SP. Z O.O. | 197663 |  |  |
| ALCON ROMANIA S.R.L. | 22018 |  |  |
| ALCON SWITZERLAND SA | 60058 |  |  |
| ALCON, FARMACEVTSKE STORITVE D.O.O. | 47706 |  |  |
| Estonia Rep. Office | 13138 |  |  |
| Latvia Rep. Office | 26660 |  |  |
| Lithuania Rep. Office | 8184 |  |  |
| Arthrex | 559718 | Arthrex | 559,718 |
| Ascensia Diabetes Care Holdings AG | 164920 | Ascensia Diabetes Care Holdings AG | 164,920 |
| B. BRAUN MEDICAL, S.A. | 880211 | B. BRAUN | 4,961,969 |
| B. Braun Melsungen AG | 1644079 |  |  |
| B. Braun SE | 1203576 |  |  |
| B. BRAUN SURGICAL, S.A. | 1234103 |  |  |
| BARNA IMPORT MEDICA, S.A. | 1107831 | BARNA IMPORT MEDICA, S.A. | 1,107,831 |
| Bausch & Lomb U.K. Limited | 6358 | Bausch & Lomb U.K. Limited | 6,358 |
| Baxter AG | 143663 | Baxter | 16,028,031 |
| Baxter AG (Serbia) | 18877 |  |  |
| Baxter B.V. | 30568 |  |  |
| Baxter Bulgaria EOOD | 8125 |  |  |
| Baxter Czech spol. s r.o. | 38763 |  |  |
| Baxter D.O.O Slovenia | 44465 |  |  |
| Baxter Deutschland GmbH | 464192 |  |  |
| Baxter Estonia OU | 1500 |  |  |
| Baxter Healthcare d.o.o. Croatia | 39030 |  |  |
| Baxter Healthcare GmbH | 273233 |  |  |
| Baxter Healthcare ltd | 139762 |  |  |
| Baxter Healthcare SA | 291576 |  |  |
| Baxter Hungary Kft. | 8546 |  |  |
| Baxter Latvia SIA | 8500 |  |  |
| Baxter Medical AB | 29167 |  |  |
| Baxter Oy | 1000 |  |  |
| Baxter Polska Sp.z.o.o. | 54838 |  |  |
| BAXTER S.P.A. | 899133 |  |  |
| Baxter Slovakia s.r.o. | 5130 |  |  |
| Baxter Turkey Renal Hizmetleri A.Ş. | 71374 |  |  |
| BAXTER, S.L. | 13450089 |  |  |
| UAB Baxter Lithuania | 6500 |  |  |
| Bayer Hispania | 1888718 | Bayer Hispania | 1,888,718 |
| BD | 966366 | BD | 966,366 |
| Beckman Coulter | 141021 | Beckman Coulter | 141,021 |
| Bio-Rad Europe GmbH | 22701 | Bio-Rad Europe GmbH | 22,701 |
| BIOMERIEUX | 2341464 | BIOMERIEUX | 2,341,464 |
| Biomet 3i Dental Iberica SLU | 608951 | Biomet 3i Dental Iberica SLU | 608,951 |
| BIOTRONIK | 2600265 | BIOTRONIK | 7,486,324 |
| BIOTRONIK ApS | 43402 |  |  |
| BIOTRONIK HUNGARIA Kereskedelmi Kft. | 80507 |  |  |
| BIOTRONIK Italia SpA | 625518 |  |  |
| BIOTRONIK Oy | 30552 |  |  |
| BIOTRONIK Schweiz AG | 3312660 |  |  |
| BIOTRONIK SE & Co. KG | 96184 |  |  |
| BIOTRONIK SPAIN S.A. | 675903 |  |  |
| BIOTRONIK U.K. LIMITED | 21332 |  |  |
| Boston Scientific | 33137326 | Boston Scientific | 33,137,326 |
| BTG plc | 153793 | BTG plc | 153,793 |
| Cardinal Health Ireland Unlimited Company | 3221314 | Cardinal Health | 3,343,307 |
| Cardinal Health Switzerland 515 GmbH | 121993 |  |  |
| Carl Zeiss Meditec Iberia | 7998571 | Carl Zeiss Meditec Iberia | 7,998,571 |
| Cepheid Europe | 4204 | Cepheid Europe | 4,204 |
| Cerner Iberia | 13000 | Cerner Iberia | 13,000 |
| Cerus Europe | 28634 | Cerus Europe | 28,634 |
| Coloplast A/S | 3395234 | Coloplast | 3,400,814 |
| Coloplast Oy | 5581 |  |  |
| ConMed Iberia S.L. | 175086 | ConMed Iberia S.L. | 175,086 |
| Convatec | 269809 | Convatec | 274,184 |
| KVTECH | 4375 |  |  |
| COOK MEDICAL | 3486182 | COOK MEDICAL | 3,486,182 |
| Corin Limited | 128576 | Corin Limited | 128,576 |
| CR Bard Inc | 886848 | CR Bard Inc | 886,848 |
| CVRx Switzerland LLC | 301714 | CVRx Switzerland LLC | 301,714 |
| DIAGNOSTICA STAGO SL | 196620 | DIAGNOSTICA STAGO SL | 196,620 |
| DiaSorin S.p.A. | 102484 | DiaSorin S.p.A. | 102,484 |
| DiaSys Diagnostic Systems GmbH | 3604 | DiaSys Diagnostic Systems GmbH | 3,604 |
| Direx | 85552 | Direx | 85,552 |
| Edwards Lifesciences | 6874399 | Edwards Lifesciences | 6,874,399 |
| Electramed Ltd. | 10000 | Electramed Ltd. | 10,000 |
| Elekta Medical SAU | 89180 | Elekta Medical SAU | 89,180 |
| Endologix International B.V. | 524154 | Endologix International B.V. | 524,154 |
| Endomagnetics Limited | 17482 | Endomagnetics Limited | 17,482 |
| Erbe Polska Sp. z o.o. | 127977 | Erbe Polska Sp. z o.o. | 127,977 |
| Essity Hygiene and Health AB | 238909 | Essity Hygiene and Health AB | 238,909 |
| Esteve Teijin Healthcare, S.L. | 72363 | Esteve Teijin Healthcare, S.L. | 72,363 |
| Exactech International Operations, AG | 542151 | Exactech International Operations, AG | 542,151 |
| FERRER INTERNACIONAL | 44799 | FERRER INTERNACIONAL | 44,799 |
| Fleetwood Healthcare Ltd | 2315 | Fleetwood Healthcare Ltd | 2,315 |
| FRESENIUS KABI ESPAÑA, S.A.U. | 272527 | FRESENIUS KABI ESPAÑA, S.A.U. | 272,527 |
| Fresenius Medical Care Deutschland GmbH | 1283099 | Fresenius Medical Care Deutschland GmbH | 1,283,099 |
| Fujirebio Europe NV | 311507 | Fujirebio Europe NV | 311,507 |
| GE Medical Systems | 1171003 | GE Medical Systems | 1,171,003 |
| Genomic Health | 51121 | Genomic Health | 51,121 |
| Globus Medical | 97401 | Globus Medical | 97,401 |
| GRIFOLS INTERNATIONAL | 224007 | GRIFOLS INTERNATIONAL | 224,007 |
| HAIN LIFESCIENCE SPAIN, S.L. | 127914 | HAIN LIFESCIENCE SPAIN, S.L. | 127,914 |
| HAMMERMED MEDICAL POLSKA SP. Z O.O. SP.K. | 615899 | HAMMERMED MEDICAL POLSKA SP. Z O.O. SP.K. | 615,899 |
| HEXACATH ESPAÑA S.L. | 171096 | HEXACATH ESPAÑA S.L. | 171,096 |
| Hollister Incorporated | 508015 | Hollister Incorporated | 508,015 |
| Hologic Austria GmbH | 11619 | Hologic | 706,345 |
| Hologic BVBA | 110000 |  |  |
| Hologic Deutschland GmbH | 6200 |  |  |
| Hologic Iberia S.L. | 135841 |  |  |
| Hologic Italia S.R.L. | 152434 |  |  |
| Hologic Ltd. | 213661 |  |  |
| Emsor, SL | 76592 |  |  |
| Illumina, Inc. | 47997 | Illumina, Inc. | 47,997 |
| Insulet International Ltd | 40158 | Insulet International Ltd | 40,158 |
| Integra LifeSciences Corporation | 569587 | Integra LifeSciences Corporation | 569,587 |
| Intuitive Surgical Sarl | 277713 | Intuitive Surgical Sarl | 277,713 |
| Johnson & Johnson Medical | 182,689,416 | Johnson & Johnson Medical | 182,689,416 |
| LABORATORIOS URGO S.L. - URGO MEDICAL | 99301 | LABORATORIOS URGO S.L. - URGO MEDICAL | 99,301 |
| Laker Pharmaceuticals Ltd | 50000 | Laker Pharmaceuticals Ltd | 50,000 |
| LEXEL SL | 1496 | LEXEL SL | 1,496 |
| Li | 79239 | Li | 79,239 |
| Linde Médica | 34696 | Linde Médica | 37,670 |
| LINDE MEDICA, S.L.U. | 2974 |  |  |
| LivaNova PLC | 1224926 | LivaNova PLC | 1,224,926 |
| MACO SPANIA, S.L | 31818 | MACO SPANIA, S.L | 31,818 |
| Medacta International | 319362 | Medacta International | 319,362 |
| Medela AG | 1000 | Medela | 26,877 |
| Medela Medizintechnik GmbH & Co. Handels KG | 1000 |  |  |
| Medela Polska Sp. z o.o. | 1629 |  |  |
| Medela UK Ltd. | 3762 |  |  |
| OOO Medela | 19486 |  |  |
| Medical Measurement Systems BV | 9760 | Medical Measurement Systems BV | 9,760 |
| Medline International BV | 478235 | Medline International BV | 478,235 |
| Medtronic International Trading Sàrl | 23754792 | Medtronic International Trading Sàrl | 23,754,792 |
| MENARINI DIAGNÓSTICOS, S.A. | 218005 | MENARINI DIAGNÓSTICOS, S.A. | 218,005 |
| Merit Medical Nederland BV | 13600 | Merit Medical Nederland BV | 13,600 |
| MHC Belgium (BeNELux) | 24654 | Mölnlycke Health Care | 375,117 |
| MHC Croatia | 3000 |  |  |
| MHC Cz. Republic | 31702 |  |  |
| MHC Germany | 8 |  |  |
| MHC Hungary | 40812 |  |  |
| MHC Italy | 29500 |  |  |
| MHC Poland | 8981 |  |  |
| MHC Sweden | 16053 |  |  |
| Mölnlycke Health Care | 178950 |  |  |
| Mölnlycke Health Care Russia | 31487 |  |  |
| Molnlycke Health Care s.r.o. | 2500 |  |  |
| Mölnlycke Health Care SEE | 7470 |  |  |
| MicroPort CRM SARL | 283706 | MicroPort | 323,706 |
| MicroPort Orthopedics Inc. | 40000 |  |  |
| MyNewCompany | 10611 | MyNewCompany | 10,611 |
| Nevro | 372691 | Nevro | 372,691 |
| nueva prueba | 1000 | nueva prueba | 1,000 |
| NuVasive Inc. | 1292442 | NuVasive Inc. | 1,292,442 |
| Olympus Czech Group, s.r.o., člen koncernu (OCG) | 86575 | Olympus | 1,124,468 |
| Olympus Deutschland GmbH | 22000 |  |  |
| Olympus Europa SE & Co. KG | 757779 |  |  |
| Olympus Iberia S.A.U. | 23303 |  |  |
| Olympus KeyMed | 37671 |  |  |
| Olympus Moscow LLC. | 177305 |  |  |
| Olympus Sverige AB | 19835 |  |  |
| Ortho Clinical Diagnostics | 816771 | Ortho Clinical Diagnostics | 816,771 |
| Orthofix | 150884 | Orthofix | 349,058 |
| Orthofix International N.V. | 198174 |  |  |
| PALEX MEDICAL SAU | 833181 | PALEX MEDICAL SAU | 833,181 |
| PAUL HARTMANN ADRIATIC D.O.O. | 13181 | PAUL HARTMANN | 403,875 |
| PAUL HARTMANN AG | 55647 |  |  |
| PAUL HARTMANN D.O.O. | 11832 |  |  |
| Paul Hartmann Ges.m.b.H. | 315 |  |  |
| PAUL HARTMANN Polska Sp. z o.o. | 46481 |  |  |
| PAUL HARTMANN Romania | 8445 |  |  |
| PAUL HARTMANN S.p.A. | 70054 |  |  |
| PAUL HARTMANN UK | 2848 |  |  |
| Laboratorios HARTMANN S.A | 16830 |  |  |
| IVF HARTMANN AG | 160899 |  |  |
| HARTMANN-Rico Hungária Kft | 11135 |  |  |
| HARTMANN RICO s.r.o. | 4000 |  |  |
| BODE Chemie GmbH | 2208 |  |  |
| PEI Surgical | 169724 | PEI Surgical | 169,724 |
| Radiometer | 110952 | Radiometer | 117,211 |
| Radiometer Ibérica SL | 6259 |  |  |
| Renishaw Neuro Solutions | 1255 | Renishaw Neuro Solutions | 1,255 |
| RESMED | 1500 | RESMED | 61,206 |
| RESMED EPN ITALY | 13350 |  |  |
| RESMED EPN LTD SUCURSAL EN ESPAñA | 32604 |  |  |
| ResMed Nederland BV | 1650 |  |  |
| Resmed Norway A/S | 4060 |  |  |
| ResMed UK | 8042 |  |  |
| Roche Diabetes Care Austria | 35359 | Roche | 7,367,869 |
| Roche Diabetes Care Deutschland GmbH | 10000 |  |  |
| Roche Diabetes Care Italy | 24572 |  |  |
| Roche Diabetes Care Russia | 122543 |  |  |
| Roche Diagnostics | 6014827 |  |  |
| Roche Diagnostics Austria | 12164 |  |  |
| Roche Diagnostics Croatia | 86829 |  |  |
| Roche Diagnostics Czech Republic | 27408 |  |  |
| Roche Diagnostics Deutschland GmbH | 9485 |  |  |
| Roche Diagnostics Hungary | 101604 |  |  |
| Roche Diagnostics Ireland | 600 |  |  |
| Roche Diagnostics Italy | 138571 |  |  |
| Roche Diagnostics Lithuania | 17900 |  |  |
| Roche Diagnostics Netherland | 51775 |  |  |
| Roche Diagnostics Poland | 104809 |  |  |
| Roche Diagnostics Romania | 3764 |  |  |
| Roche Diagnostics Slovenia | 84969 |  |  |
| Roche Diagnostics Spain | 129027 |  |  |
| Roche Diagnostics Switzerland | 60548 |  |  |
| Roche Diagnostics Turkey | 260512 |  |  |
| Roche Diagnostics UK Ltd. | 70603 |  |  |
| Royal Philips | 1273871 | Royal Philips | 1,273,871 |
| RS MEDICAL COMPANY S.L. | 42617 | RS MEDICAL COMPANY S.L. | 42,617 |
| Siemens Healthcare | 2651317 | Siemens Healthcare | 2,651,317 |
| Smith & Nephew Orthopaedics AG | 7596393 | Smith & Nephew Orthopaedics AG | 7,596,393 |
| Smiths Medical | 73402 | Smiths Medical | 73,402 |
| STAGO ITALIA | 224061 | STAGO ITALIA | 224,061 |
| straumanngroup | 107180 | straumanngroup | 107,180 |
| Stryker European Operations B.V. | 6312876 | Stryker European Operations B.V. | 6,312,876 |
| Sysmex Deutschland GmbH | 22550 | Sysmex | 381,314 |
| Sysmex España | 32971 |  |  |
| Sysmex España S.L. | 13042 |  |  |
| Sysmex Europe GmbH | 193903 |  |  |
| Sysmex Suisse AG | 46759 |  |  |
| Sysmex Turkey Diagnostik Sistemleri Limited Sirketi | 72089 |  |  |
| Teleflex Medical Europe | 235777 | Teleflex Medical Europe | 235,777 |
| TERUMO BCT EUROPE NV | 540036 | TERUMO | 4,419,226 |
| Terumo Europe NV | 2234934 |  |  |
| TERUMO EUROPE NV | 1644256 |  |  |
| The Binding Site Spain (Specialist Protein Company), S.L. | 71176 | The Binding Site Spain (Specialist Protein Company), S.L. | 71,176 |
| Therakos (UK) Ltd | 1430756 | Therakos (UK) Ltd | 1,430,756 |
| Thermo Fisher Scientific | 1462780 | Thermo Fisher Scientific | 1,462,780 |
| VIRCELL SPAIN SLU | 3327831 | VIRCELL SPAIN SLU | 3,327,831 |
| W.L Gore & Associates GmbH | 3926628 | W.L Gore & Associates GmbH | 3,926,628 |
| Wallac Oy | 41364 | Wallac Oy | 41,364 |
| Wellspect Healthcare | 102975 | Wellspect Healthcare | 102,975 |
| Werfen | 907409 | Werfen | 4,127,629 |
| Comesa | 167866 |  |  |
| Izasa Hospital | 887634 |  |  |
| Izasa Scientific | 2153820 |  |  |
| Nicolai | 10900 |  |  |
| Zimmer GmbH | 6971635 | Zimmer GmbH | 6,971,635 |

### Table 2. Summary of revenue, R&D and product portfolios for top 10 companies

|  | **Company** | **In Vitro Diagnostic Device and/or Medical Devices** | **Summary of product portfolios** | **2019 R&D spend**  **(million €)** | **2019 revenue or sales**  **(million €)** | **Overall payments per million revenue** |
| --- | --- | --- | --- | --- | --- | --- |
| 1 | Johnson & Johnson Medical^3^ | Medical Devices | 1. Surgery 2. Orthopaedics 3. Vision | 1,812 | 23,192 | 7,887 |
| 2 | Abbott Laboratories^4^ | In Vitro Diagnostic Device & Medical Devices | 1. Core Laboratory 2. Rhythm Management 3. Electrophysiology | 2,180* | 17,822 | 2,449 |
| 3 | Boston Scientific^5^ | Medical Devices | 1. Interventional Cardiology 2. Cardiac Rhythm Management 3. Endoscopy | 1,049* | 9,517 | 3,482 |
| 4 | Medtronic International Trading Sàrl^6^ | Medical Devices | 1. Cardiac Rhythm & Heart Failure 2. Surgical Innovations 3. Coronary and Structural Heart | 2,081 | 27,295 | 870 |
| 5 | Baxter^7^ | Medical Devices | 1. Renal Care^t^ 2. Medication Delivery 3. Advanced Surgery^t^ | 531* | 10,149* | 1,579 |
| 6 | Carl Zeiss Meditec Iberia^8^ | N.A. | N.A. | 630* | 5,742* | 1,393 |
| 7 | Smith & Nephew Orthopaedics AG^9^ | Medical Devices | 1. Orthopaedics 2. Sports Medicine & ENT 3. Advanced Wound Management | 261 | 4,590 | 1,655 |
| 8 | BIOTRONIK | Medical Devices | N.A. | N.A. | N.A. |  |
| 9 | Roche^10^ | In Vitro Diagnostic Device & Medical Devices | 1. Centralised and Point of Care Solutions 2. Molecular Diagnostics 3. Diabetes Care | 1,396 | 11,719 | 629 |
| 10 | Zimmer GmbH^11^ | Medical Devices | 1. Knees 2. Hips 3. Surgical, sports medicine, biologics, foot and ankle, extremities and trauma products | 401 | 7,130 | 978 |

*This figure represents the company’s overall revenue or R&D spend as separate figures were not provided for medical devices only

^t^it is not clear whether or not this area includes pharmacological products

### Box 5. Changes to Accessibility, Availability and Quality Proforma

A new section was added; *availability,* which contains three measures. Firstly, the extent to which different recipient categories are included. Secondly, whether all medical device companies in the territory are included. Thirdly, the extent to which different payment categories are included. Also, three measures were added to the accessibility section. Firstly, whether disclosures are removed after a certain number of years. Secondly, the availability of methodology notes. Methodology notes provide details of how each medical device company recognises, classifies and reports relevant payments. And thirdly, whether all relevant terminology is defined. Two measures were modified *Spectrum of disclosed characteristics* and *Database searchability*. For searchability, to gain a high rating in the new proforma, the database has to have advanced search capabilities, a medium rating requires limited search capabilities. In the previous proforma, limited search capabilities were sufficient for a high rating. For *Spectrum of disclosed characteristics*, to achieve a high rating in this modified proforma, the database has to include all characteristics from the US Open Payments database. Whereas in the previous proforma the database had to include all characteristics from the trade body’s disclosure template.

### Table 3. Accessibility, availability and quality of transparentmedtech.eu (relevant ratings underlined and written in bold)

| **Accessibility** |  | Higher accessibility |  | Lower accessibility | Rationale |
| --- | --- | --- | --- | --- | --- |
| Database format | How is the database published (ie, PDF, XLS, CSV, webpage)? | **Webpage, XLS or**  **CSV** | Readable PDFs | Image-based PDFs | Database is available on a website |
| Database structure | Does the data from all companies follow a single template consistently? | **Yes** | N/A | No | All entries appear in the same table with common headings, and further information on each entry uses the same template |
| Database searchability | Can the database be searched? If so, can database searches be carried out without data users providing any additional information? | Advanced search available | **Limited search available** | No search available | A search bar is available on the website, however no advanced features are available |
| Customisable summary statistics | Does the database offer users the possibility of generating real-time, dynamic data summaries based on selected database characteristics? | Yes | N/A | **No** | No summary statistics are available |
| Unique identifiers | Do reported donors (medical device companies) and recipients (healthcare organisations) have unique identifiers? | All donors and recipients | **Some donors or recipients** | No unique identifiers | Some donors and recipients have unique identifiers but there are many examples of recipient entries with no unique identifier provided |
| Downloadability | Can the database be downloaded (eg, as a single CSV or XLS file) for further analysis? | Yes | N/A | **No** | The database cannot be downloaded |
| Methodology notes | Are notes describing the methodology for each company available | Available online | **Available on request** | Not available | The code of conduct specifies: ‘methodology note shall be made available upon request by an interested party’ |
| Year limit | Are disclosures removed after a certain number of years | No | N/A | **Yes** | 2017 data is no longer available on the site |
| Definitions | Is all relevant terminology defined | Clear and comprehensive definitions are provided for all relevant terminology (e.g. healthcare organisation, payment categories) | **Definitions are provided for some relevant terminology** | No definitions are provided for relevant terminology | Definitions are not provided for *Support to Educational Events* and *Other Educational Grants*, only examples are provided |
| **Quality** |  | Higher quality |  | Lower quality |  |
| Spectrum of disclosed characteristics | What characteristics are included in relation to donors, recipients and payments? | All characteristics from the US Open Payments database covered | **Some of the characteristics from the US Open Payments database covered** | None of the characteristics from the US Open Payments database covered | Examples of characteristics not included: therapeutic area payment is related to; Recipient type (e.g. hospital) |
| Aggregation of payments | Are payments itemised (ie, all payments have separate entries) or are they aggregated on an annual basis (eg, per recipient and/or payment category)? | All payments itemised | **Some payments itemised, others aggregated** | All payments aggregated | According to guidelines, all payments should be aggregated but there are some examples of itemised payments (different entries for payments from one medical device company to one HCO in a single year and in a single area e.g. *Other Educational Grants*) |
| Inclusion of taxes | Is it clear whether payments are reported inclusive or exclusive of any taxes, such as VAT? | **Single rule for all companies and payments** | No single rule, each company sets its own rules for VAT reporting which are published separately from payment disclosures | Rules around tax reporting are unclear | Guidelines stipulate that VAT should be excluded from all reported payments |
| **Availability** |  | Higher availability |  | Lower availability |  |
| Breadth of recipients | Are patient organisations, healthcare organisations, and healthcare professionals included | All of the following groups of recipients are included: Patient organisations, healthcare organisations, healthcare professionals | N/A | **Some of the following groups of recipients are not included: Patient organisations, healthcare organisations, healthcare professionals** | According to guidelines, patient organisations, and healthcare professionals are not included |
| Breadth of donors | Are all medical device companies in the territory included | all medical device companies in the territory are included | N/A | **Some of the medical device companies in the territory are not included** | According to a BMJ article, less than 10% of UK medical device companies are members of the UK branch of MedTech Europe^12^ |
| Breadth of payment areas | Are the payment areas comprehensive | All of the following types of payments included: education, research, consulting, gifts, charitable donations, 2  ownership and investment interest | N/A | **Some of the following types of payments are not included: education, research, consulting, gifts, charitable donations, ownership and investment interest** | The following types of payments are not included: research, consulting, gifts, charitable donations, ownership and investment interests |

### Table 4. Payments for non-member countries’ HCOs and HCOs of member countries with superseding national regulations

| **Country** | **Total value of payments (€) 2017** | **Total value of payments (€) 2018** | **Total value of payments (€) 2019** | **Total value of payments (€) 2017-2019** |
| --- | --- | --- | --- | --- |
| *Member countries with superseding national legislation* |  |  |  |  |
| Belgium | 40,825 | 1,377,136 | 99,606 | 1,517,567 |
| France | 313,225 | 1,594,292 | 269,092 | 2,176,609 |
| Netherlands | 7,005,846 | 7,232,703 | 7,233,347 | 21,471,896 |
| Portugal | 22,261 | 79,781 | 17,346 | 119,387 |
| *Non-Member Countries* |  |  |  |  |
| Armenia | 0 | 0 | 4,000 | 4,000 |
| Australia | 8,852 | 0 | 0 | 8,852 |
| Bosnia & Herzogovina | 1,830 | 13,590 | 0 | 15,420 |
| Cameroon | 0 | 0 | 1,500 | 1,500 |
| Chile | 0 | 0 | 7,659 | 7,659 |
| Egypt | 0 | 0 | 11,857 | 11,857 |
| Georgia | 0 | 0 | 480 | 480 |
| Israel | 0 | 0 | 3,000 | 3,000 |
| Japan | 0 | 0 | 447 | 447 |
| Kazakhstan | 0 | 0 | 2,760 | 2,760 |
| Kuwait | 0 | 0 | 2,274 | 2,274 |
| Lebanon | 0 | 0 | 7,226 | 7,226 |
| Monaco | 0 | 1,860 | 45,000 | 46,860 |
| Niger | 0 | 0 | 30,500 | 30,500 |
| Serbia | 64,565 | 50,000 | 20,883 | 135,448 |
| South Africa | 0 | 0 | 18,252 | 18,252 |
| Sudan | 0 | 0 | 2,500 | 2,500 |
| Tunisia | 0 | 0 | 2,480 | 2,480 |
| Ukraine | 0 | 0 | 73,155 | 73,155 |
| United Arab Emirates | 0 | 0 | 10,914 | 10,914 |
| United States | 0 | 0 | 46,039 | 46,039 |
| Total | 7,457,404 | 10,349,362 | 7,910,317 | 25,717,082 |

### Table 5. Payments in member countries^*^ without superseding national legislation

|  | **Total value of payments (€)** | | | |  |  |
| --- | --- | --- | --- | --- | --- | --- |
| **Country** | **2017** | **2018** | **2019** | **2017-2019** | **% increase from 2017 to 2019** | **€ per 1,000 population (2017-2019)** |
| Austria | 1,883,884 | 3,667,607 | 3,597,512 | 9,149,003 | 91.0% | 1,033 |
| Bulgaria | 9,523 | 54,775 | 80,678 | 144,976 | 747.2% | 21 |
| Croatia | 522,403 | 1,095,836 | 798,755 | 2,416,995 | 52.9% | 593 |
| Cyprus | 0 | 14,659 | 0 | 14,659 | 0% | 4 |
| Czechia | 222,247 | 1,730,129 | 1,371,080 | 3,323,456 | 516.9% | 312 |
| Denmark | 396,613 | 703,354 | 650,711 | 1,750,678 | 64.1% | 302 |
| Estonia | 4,800 | 139,165 | 250,567 | 394,532 | 5120.1% | 298 |
| Finland | 148,823 | 341,661 | 366,209 | 856,694 | 146.1% | 155 |
| Germany | 3,954,334 | 8,328,356 | 8,740,666 | 21,023,355 | 121.0% | 253 |
| Greece | 636,631 | 1,204,514 | 1,680,122 | 3,521,268 | 163.9% | 328 |
| Hungary | 217,738 | 702,377 | 665,838 | 1,585,953 | 205.8% | 162 |
| Iceland | 16,000 | 9,911 | 1,965 | 27,876 | -87.7% | 78 |
| Ireland | 179,561 | 710,025 | 1,223,359 | 2,112,945 | 581.3% | 441 |
| Italy | 4,478,843 | 11,082,551 | 14,913,356 | 30,474,751 | 233.0% | 505 |
| Latvia | 39,768 | 206,149 | 157,540 | 403,456 | 296.1% | 210 |
| Lithuania | 74,140 | 167,905 | 180,462 | 422,506 | 143.4% | 151 |
| Luxembourg | 492,386 | 274,688 | 551,708 | 1,318,782 | 12.0% | 2,148 |
| Norway | 119,355 | 123,503 | 31,584 | 274,442 | -73.5% | 52 |
| Poland | 1,021,517 | 2,417,408 | 2,108,281 | 5,547,206 | 106.4% | 146 |
| Romania | 65,887 | 151,030 | 159,738 | 376,655 | 142.4% | 19 |
| Russia | 760,324 | 1,062,532 | 2,490,866 | 4,313,722 | 227.6% | 30 |
| Slovakia | 21,526 | 345,742 | 303,114 | 670,382 | 1308.1% | 123 |
| Slovenia | 281,295 | 336,248 | 454,575 | 1,072,118 | 61.6% | 515 |
| Spain | 8,864,272 | 27,154,209 | 49,150,427 | 85,168,908 | 454.5% | 1,815 |
| Sweden | 203,562 | 469,939 | 206,193 | 879,694 | 1.3% | 86 |
| Switzerland | 51,655,608 | 62,963,113 | 61,876,963 | 176,495,685 | 19.8% | 20,661 |
| Turkey | 1,243,105 | 1,895,111 | 1,357,283 | 4,495,499 | 9.2% | 55 |
| UK | 6,318,120 | 16,154,799 | 14,134,432 | 36,607,351 | 123.7% | 549 |
| Total | 83,832,265 | 143,507,296 | 167,503,984 | 394,843,547 | 99.8% | 607 |

*Payments were made to HCOs registered in the above countries

### Box 6. Results of content analysis inductively examining other issues with the database

The content analysis revealed several other issues with the database beyond what was assessed in the accessibility, availability and quality proforma. MedTech Europe disclosure guidance is clear that transparentmedtech.eu should be used for payments to HCOs, and patient organisations are explicitly excluded. Still, the database includes some patient organisations (e.g. the Slovenian Kidney Patient Association). Also, there are examples of entries that appear to be the names of individuals rather than HCOs.

The name of some HCOs is not provided. A notable entry in Table 5 is ‘European HCO and PCO’ which is in receipt of the third largest amount of any HCO. This is likely to be an aggregation of several payments to different healthcare organisations and professional conference organisers by B. Braun. Another example is ‘Switzerland Aggregated Disclosure (Entities That Have Not Provided Consent)’ which is assigned a total payment of €155,878 from Johnson and Johnson Medical. MedTech Europe guidance stipulates that a medical device company’s payments for a given area, in a given year should be aggregated for an individual healthcare organisation. There are several instances where this is not adhered to. For example, Roche disclosed several different payments to Szpital Uniwersytecki in 2019.

There are several cases of payments disclosed on the transparentmedtech.eu website but not the respective mandatory disclosure system. For example on transparentmedtech.eu, Terumo registered a €550 payment in 2019 to the Belgian registered HCO, the International Diabetes Federation; this payment does not appear to have been registered on the Belgian mandatory disclosure website.^13^ Also, a €1287.82 payment from Werfen to the Portuguese Hospital Do Divino Espirito Sa in 2018 and a €1455 payment from Werfen to the Portuguese Laboratorio De Anatomia Patolo in 2018 do not appear on the Portuguese website.^14^ A €300 payment from Smiths Medical to the French Société De Pneumologie De Langue Française in 2018 was not reported on the French transparency website.^15^ Also, payments to HCOs in a large number of countries who are not subject to the code (e.g. Israel, Japan, Cameroon, Chile, Egypt) are reported on the website.
